# Epidemiological and socio-economic characteristics of the COVID-19 spring outbreak in Quebec, Canada: A population-based study

**DOI:** 10.1101/2020.08.26.20182675

**Authors:** Rodolphe Jantzen, Nolwenn Noisel, Sophie Camilleri-Broët, Catherine Labbé, Thibault de Malliard, Yves Payette, Philippe Broët

## Abstract

**Background:** By mid-July 2020, more than 108,000 COVID-19 cases had been diagnosed in Canada with more than half in the province of Quebec. To be prepared for a potential second wave of COVID-19 in the fall, it seems of utmost importance to analyze the epidemiological and socio-economic characteristics of the spring outbreak in the population.

**Method:** We conducted an online survey of the participants of the CARTaGENE population-based cohort, composed of middle-aged and older adults. We collected information on socio-demographic, lifestyle, health condition, COVID-related symptoms and COVID-19 testing. We studied the association between these factors and two outcomes: the status of having been tested for SARS-CoV-2 and the status of having received a positive test when having been tested. These associations were evaluated with univariate and multivariate analyzes using a hybrid tree-based regression model.

**Results:** Among the 8,129 respondents from the CARTaGENE cohort, 649 were tested for COVID-19 and 41 were positive. Medical workers and individuals having a contact with a COVID-19 patient had the highest probabilities of being tested (32% and 42.4%, respectively) and of being positive (17.2% and 13.0%, respectively) among those tested. 7.6% of the participants declared that they have experienced at least one of the four COVID-related symptoms chosen by the Public Health authorities (fever, cough, dyspnea, anosmia) but were not tested. Results from the tree-based model analyzes adjusted on exposure factors show that the combination of dyspnea, dry cough and fever was highly associated with being tested whereas anosmia, fever, and headache were the most discriminant factors for having a positive test among those tested. During the spring outbreak, more than one third of the participants have experienced a decrease in access to health services. There were sex and age differences in the socio-economic and emotional impacts of the pandemic.

**Conclusion:** We have shown some discrepancies between the symptoms associated with being tested and being positive. In particular, the anosmia is a major discriminant symptom for positivity whereas ear-nose-throat symptoms seem not to be COVID-related. The results also emphasize the need of increasing the accessibility of testing for the general population.

## Background

By mid-July 2020, more than 108,000 COVID-19 cases had been diagnosed in Canada with more than half in the province of Quebec [1]. The province of Quebec confirmed its first case of COVID-19 on February 27. With only seventeen confirmed cases, the state of emergency was declared by the Quebec government on March the 13th with a shutdown of daycare facilities, primary and secondary schools, CEGEPs and Universities followed by a more extensive shutdown of restaurants and bars, indoor sport facilities and non-essential economic activities. In early March, most cases could be traced to infected travelers returning to Canada or to close contact with those travelers. By March 23, according to the Public Health Agency of Canada, nearly half of Canadian COVID-19 cases have been acquired through community spread [2]. In mid-July, more than 56,000 COVID-19 postive cases have been confirmed in the province of Quebec with nearly half in the Montreal metropolitan area and around one-third of the confirmed cases have been diagnosed among middle-aged and older adults. As in many countries over the world, the COVID-19 testing strategy in Canada has evolved over the first wave, taking into account each province’s specific situation.

In order to be prepared for a potential second wave of COVID-19 in the fall, it seems of utmost importance to analyze the epidemiological and socio-economic characteristics of the spring outbreak in the population, together with the results of the public health policies that have been implemented. Such information can provide useful knowledge for planning public health strategies for the next coming months.

Despite the impressive number of publications about COVID-19 infection, most of them are hospital-based series, [3, 4, 5, 6] to name a few, while population-based studies are scarce [7, 8, 9]. However, population-based cohorts that have been established before the pandemic may yield unbiased estimates of the characteristics and consequences of the pandemic in the general population. It may also provide useful information about the effectiveness of public health interventions such as testing strategies. In this context, the existing population-based cohort CARTaGENE (CaG), composed of middle-aged and older adults, which was established before the COVID-19 outbreak, offers a unique opportunity to analyze the characteristics and consequences of the outbreak among a population that seems to be at greatest risk.

A large survey (hereinafter referred to as CaG COVID-19) was launched in early June. An online questionnaire was sent to the participants of the CaG cohort for collecting information about COVID-19-related symptoms, diagnosis, comorbidities and social impacts of the spring outbreak. The aim of this survey was to analyze the demographic, clinical and socio-economic characteristics associated with the COVID-19 pandemic in Quebec in spring 2020. We also investigated the risk exposure, pre-existing medical conditions and clinical features that led to being tested and to have a positive result among those who have been tested.

## Methods

### CARTaGENE population-based cohort

CARTaGENE is a population-based cohort composed of more than 43,000 Quebec residents aged between 40 and 69 years at recruitment [10]. Original survey design was defined by gender, age groups and forward sortation area (FSA - defined by the first 3-digit postal codes). Participants have been recruited during two phases (Phase A: 2009-2010 (n = 23,000) and Phase B: 2013-2014 (n = 20,000)) in metropolitan areas where nearly 70% of Quebecers live and prospective follow-ups are conducted on a regular basis. With a rich collection of data including phenotyping and biological data, CaG is the largest ongoing prospective population cohort and biobank in Quebec, Canada. The collected data includes a self-administered socio-demographic and lifestyle questionnaire (Phase A and B), an interviewer-administered health questionnaire (Phase A and B); non-invasive physical measurements (Phase A) and biospecimen collection (blood (Phase A and B) and urine (Phase A)). More than 30,000 individuals have provided blood samples. The lifestyle questionnaires cover topics such as socio-demographic factors, lifestyle, mental state, psychosocial environment, personal and family history of disease, health care utilization, medication use, reproductive health and history and declared health conditions. A cohort-wide follow-up has been carried out in 2018 aiming at updating lifestyle and health information 5 to 10 years after the baseline data. CaG is part of the Canadian Partnership for Tomorrow’s Health (CanPath, former called CPTP) which is the Canada’s largest population health research platform [11].

### CaG COVID-19 online questionnaire

For this study, we have developed a specific online questionnaire [12] for collecting updated information on basic demographic variables (age, sex, household composition and postal code), COVID-19 testing, test dates and results, whether participants suspect they had an undiagnosed COVID-19 infection based on symptoms (cough, shortness of breath, fever, etc.). We also collected information on exposure status: health care or essential workers, contact with someone who has been diagnosed with COVID-19, international travel. Those with confirmed COVID-19 infection were asked if they were hospitalized along with details related to their care, treatment and outcome of the hospitalization. In addition, participants were asked to report existing chronic health conditions and medication use. Finally, they were asked about the psychosocial and socio-economic impacts of the pandemic on their lives. This online questionnaire was unfolded in close collaboration with the CanPath consortium for allowing future inter-provincial comparisons as well as other national and international research initiatives. A weblink to the consent and questionnaire was sent by email to all the CARTaGENE participants with a valid email address, that is 33,019. Initial invitation was followed by up to 3 emails reminders. The digital invitations were sent early in June 2020 and were valid for 4 weeks. Survey closed early July 2020.

### Investigated variables and outcomes

A positive exposure status was defined as an individual being either medical or essential workers, having been in contact with a COVID-19 positive patient or coming back from international travel. A medical worker was defined as either a physician, nurse, hospital/aged-care facility employee, first responder, pharmacist with patient exposure. An essential worker was defined as either a grocery store attendant, public transit, police, fireman. A contact with a COVID-19 positive individual was defined as being in the same room as a person who was told by a health professional that he or she has COVID-19. International travel was considered as a travel outside Canada returning after the 1st of February.

Pre-existing conditions were defined as medical condition currently treated among the following list: cardiovascular diseases (HBP, coronary artery disease,…), autoimmune diseases, diabetes, infectious diseases, gastro-intestinal and liver diseases, cancers, renal diseases.

The following list of symptoms were considered in the questionnaire: dry cough, wet cough, shortness of breath (dyspnea), fever (≥ 38°C), shivering, fatigue, runny nose, sinus pain (sinusitis), ear pain (otitis), sore throat, hoarseness, loss of taste (ageusia), loss of smell (anosmia), diarrhea, nausea, vomiting, loss of appetite, headache, general muscle aches and pains. The possible responses were: none, mild, moderate or severe.

For the association studies, the following symptoms were considered as present only when being severe: fatigue, loss of appetite, sinus pain, ear pain, sore throat, hoarseness, headache, muscle pain, diarrhea.

Hereafter, we will refer to the symptom-based case definition chosen by the Quebec Public Health authorities with at least one of the four major symptoms: fever (≥ 38°C), a new cough or a cough that gets worse, difficulty of breathing, anosmia with or without ageusia [13].

Two outcomes were studied. The first was the status of having been tested for SARS-CoV-2 (rt-PCR). The second was the status of having received a positive test when having been tested. Confirmed COVID-19 infection was defined as at least one positive result test. If multiple tests were performed, we considered the date of the first positive one.

### Statistical methods

#### Descriptive and univariate analyzes

The participants’ baseline characteristics have been reported and tested: demographic (age, sex, geographical location, level of education, financial resources, household income, dwelling type), health history (high blood pressure, cardio-vascular disease, pulmonary disease (chronic obstructive pulmonary disease (COPD), asthma,…), cancer, diabetes, autoimmune diseases, mental health, body mass index (BMI)), lifestyle behaviors (smoking status, alcohol intake,…).

Mean (or median) with standard deviations (or interquartile ranges) were reported for continuous outcomes. Frequencies with 95% confidence intervals were reported for qualitative variables. For comparing means between groups, we used an ANOVA test. For categorical variables, we used chi-squared test or exact test if needed.

For the univariate association analyzes (between factors and studied outcomes), we used a chi-square test (qualitative) or a logistic regression model (quantitative). For each hypothesis test, we reported the p-values. Moreover, in order to address the multiple testing problem, we also indicated those that are still significant for a FDR (the expected proportion of false discoveries among all discoveries) controlled at a level of 1% using the Benjamini-Hochberg procedure [14].

#### Multivariate analyzes

For the multivariate analyzes of exposure and risk factors, we used a multiple logistic regression model.

For the multivariate analyzes of symptoms associated with the outcomes, we had to cope with complex interplay between clinical symptoms. Thus, we considered a Generalized Partially Linear Tree-based Regression (GPLTR) model. GPLTR models represent a class of semi-parametric regression models that integrate the advantages of generalized linear regression and tree-structure models [15]. The linear part is used to model the main effects of confounding variables (e.g. exposure) while the nonparametric tree part is used to address potential collinearity and interactions between explanatory variables (e.g. clinical symptoms). This tree-based model provides a classification of individuals in homogeneous groups in terms of risk for the event of interest and identifies relevant combination of explanatory variables. The optimal GPLTR tree is selected using a penalized maximum likelihood method with the Bayesian information criterion (BIC) [15].

Regression trees are prone to instability, especially when dealing with a low number of outcomes, making variable selection somewhat precarious. Thus, for the an-alyzis of the positive status outcome, we constructed multiple trees using a bagging approach, which provides a way to assess the relevance of each variable across the set of trees using variables’ importance measures. These later results provide us some arguments regarding the reliability of the selected optimal GPLTR tree. We reported the depth deviance importance score (DDIS) of each symptom that is computed for each GPLTR model as the sum of the values of the deviance at each split based on this variable, weighted by the location of the split in the tree. These scores are summed across the set of trees, and normalized to take values between 0 and 1, with the sum of all scores equal to 1. A set of 300 GPLTR models was done and we reported the symptoms as ranked by the DDIS.

We also evaluated in our dataset the discriminative power of the Menni et al. logistic-based predictive model [9] which is based on age, sex, loss of smell and taste, fatigue, persistent cough and loss of appetite. We reported the area under the ROC curve (AUC) together with the sensitivity and specificity applying the 50% probability threshold (as proposed by Menni et al. and corresponding to the Bayes rule).

Statistical analyzes were performed using R software [16]. Regression tree analyzes were performed with the ‘GPLTR’ R package [15].

#### Estimation of the probability of being positive when experiencing a symptom

Since the tested participants are selected based on the symptoms being hypothesized to be associated with a positive test, the tested participants constitute a non-random set (ascertainment bias). Thus, it is not possible to estimate directly the probability of being infected given that the person has experienced a particular symptom since non-selected (untested) individuals are unobserved. We can only estimate the probability of being positive given that the person has experienced a particular symptom and has been tested.

Instead, we propose to report the minimum, mean and maximum values that these probabilities can take using the law of total probability. More precisely, we know that the probability of being positive given that the person has experienced a particular symptom (*P*(+|*S*)) can be expressed as:

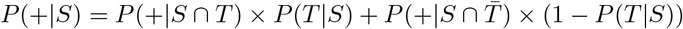

where *P*(*+|S ∩ T*) (resp. 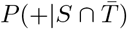 are the probabilities of being positive when the symptom is present and he/she has been tested (resp. not tested) and P(T|S) is the probability of being tested when having the symptom.

From our data, *P*(+|*S∩T*) and *P*(*T*|*S*) could be directly estimated while 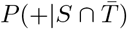 could not. However, we know that this probability ranges from zero (complete dependence) to *P*(+|*S ∩ T*) (independence between positivity and test).

Thus, we calculated the minimum, mean and maximum values that *P*(+|*S*) can attain for each symptom.

## Results

### Characteristics of CaG COVID-19 survey respondents

#### Survey respondents

Among the 43,038 participants of the CARTaGENE cohort, we sent the questionnaire to 33,019 individuals. As compared to the non-respondents (n=24,882), the respondents (n=8,137) were slightly older (median age of 62.9 vs. 60.6, *p* < 0.001), composed of more women (58.8% vs. 53.9%, *p* < 0.001), had a higher education level (58.3% vs. 44.8%, *p* < 0.001) and were more often living in Montreal (68% vs. 65.9%, *p* < 0.001). The same results were observed when comparing the individuals who answered the questionnaire to the whole CARTaGENE cohort.

Of the 8,137 respondents, 8,129 had responded to the questions regarding COVID-19 testing (“have you been tested for COVID-19?” and “What was the result of your 1st COVID-19 test?”). Hereafter in this article, we analyzed this set of 8,129 respondents.

Both the proportion of tested participants for COVID-19 and positive individuals among those being tested are consistent with those reported in Quebec at the closure of our survey (tested: CaG 8.0% [5.8-10.1] versus Quebec 7.3%; positive individuals: CaG 6.3% [4.4-8.2] versus Quebec 7.4%). We also observed a higher proportion of women among the positive individuals (CaG: 59.8% [56.0-63.6] versus Quebec 53.8%).

Among the 4,855 individuals who experienced at least one COVID-related symptom (as defined above), 1,022 declared that their first action was to call or consult their family doctor (39.3%), to call the 811 or a dedicated coronavirus hotline (24.6%), to go to the pharmacy (24.6%), to the hospital emergency room (6.4%) or to a COVID-19 screening clinic (5.1%). Among the 1,522 individuals who tried to contact a healthcare professional, 31.3% (476) were not able to reach someone on the phone or to see someone because of too much waiting.

Among the 288 participants who experienced a COVID-19 contact, the majority was at work (123), in a health care facility (69) or at home (54). In the later case, 13 COVID-19 contacts were the spouse or partner. 56.2% (162/288) were not tested.

#### Tested respondents

Among the respondents, 649 (8.0%) have been tested for COVID-19 and 41 (6.3%) were declared positive. Among these tested individuals, 548 had one test, 55 had two tests, 20 had three tests and 26 had four tests. The average time to receive a result was 3 days [IQR: 0-4 days].

When looking to the number of tests performed per day, we can see a low number of tested participants during April with a maximum at the end of May. This trend reflects the shortage faced by the province in early April followed by a rebound by end of May. In our series, the highest number of tests per day was May 22, consistent with the entire province [17] (Supplementary Figure S1).

Among the 649 tested individuals, 134 were medical workers, 123 were in contact with a COVID-19 positive individual and 69 had both risk exposures. 47 individuals were essential workers including 28 without other risk exposure. 161 individuals declared an international travel, including 117 having no other risk exposure. Among the 309 tested individuals with no declared risk exposure, 74 declared no COVID-related symptom with 41 patients having a chronic pre-existing medical condition (mainly cardio-vascular or respiratory disease).

The proportion of positive cases varied widely across risk exposure status: 17.2% (23/134) for the medical workers, 4.2% (2/47) for essential workers, 13.0% (7/54) for people in contact with a COVID-19 positive individual without professional exposure, 4.3% (5/117) for people having declared an international travel without other exposure risk and 1.9% (4/214) for people having one of the major symptoms but without known exposure risk. None of the 74 patients without risk exposure or COVID-related symptoms were positive.

There were 619 individuals who declared that they have experienced at least one of the four COVID-related symptoms chosen by the Quebec Public Health authorities but were not tested. Among them 3.4% were medical worker, 3.1% had a contact with a COVID-19 patient. Among those having none of these later exposure factors, 20.3% declared an anosmia. Moreover, the percentage decreased with the number of COVID-related symptoms from 38.1% (one symptom) to 8.4% (all the four symptoms).

#### Positive respondents

Among the 41 individuals with at least one positive test, 10 (24.4%) participants had a first negative test followed by a positive test. Twenty-three (56%) were working as a medical worker and 2 (4.9%) as an essential worker. Seven (17.0%) were in contact with a COVID-19 positive individual without professional exposure and five (12.2%) with no other risk exposure traveled outside of Canada.

Three individuals have been hospitalized for COVID-19 infection but none of them was hospitalized in ICU. Five were treated with an experimental therapy prescribed by a clinician for COVID-19. The colchicine was the only one prescribed treatment. None of them had chloroquine/hydroxychloroquine, remdesivir, lopinavir-ritonavir or tocilizumab.

Among the positive cases, 35 (89.7%) felt completely or mostly back to normal. Eight (23%) were sick for seven days or less, 12 (34%) between eight and fourteen days, 10 (29%) between fifteen and thirty days, and 5 (14%) more than thirty days. The two individuals who felt not really back to normal were at 30 and 43 days of symptoms. They had 7 and 15 different symptoms, respectively.

Among the positive individuals who responded to mental and emotional questions, 35/39 (89.7%) had someone to help meeting their immediate needs. 26/38 (68.4%) received help, aid or support (including from friends, family, community or government).

### Socio-economic, lifestyle and health early impacts of the COVID-19 spring 2020 outbreak

#### General and socio-economic impacts

While the household income had substantially decreased for 447 (5.8%) of all the participants, there was a moderate or major impact on the ability to meet financial obligations or essential needs (e.g. rent or mortgage payments, utilities and groceries) for 609 (8%).

Among the 1,302 individuals who regularly took public transit before March 2020, 919 (70.6%) did not continue to take public transit after March, while 297 (22.8%) took public transit less frequently. Individuals changed their transport habits because of the lockdown (923), being afraid of COVID-19 exposure (522), quarantine/in self-isolation (206) or COVID-19 symptoms (11).

Regarding the relationship with the intimate partner during the lockdown, 4,443 (76.9%) of the participants experienced no change, 954 (16.5%) declared that they became closer than before the pandemic, while 380 (6.6%) felt more distant or strained than before the pandemic. Same trends were observed with friends and colleagues who became closer in 1,266 (16.4%) and 921 (15.9%), respectively, while more distant in 742 (9.6%) and 421 (7.3%), respectively. The family relationship became more distant or strained than before the pandemic for 1,552 (20.2%) of the participants while 641 (8.3%) became closer.

#### Health impacts

During the spring outbreak, 2,982 (37.7%) of the participants have experienced a change in access to health services. Among them, 225 (4.5%) had a surgery canceled or deferred, 411 (8.3%) a medical procedure canceled or deferred, 289 (5.8%) a treatment canceled or deferred and 711 (14.4%) experienced a delay for seeing a healthcare professional about an preexisting problem and 344 (6.9%) about a new problem. However, 2,157 (43.7%) have used virtual appointments with their health care provider.

Among the 100 individuals who have initiated new use of mental health services, 48 did for anxiety, 35 for depression and 39 for stress. Overall, the mental/emotional health at the time of completing the questionnaire was good/excellent for 5,370 (69.2%) participants.

Over the last 2 weeks, some individuals have been bothered more than half of the days or nearly everyday by the following problems: trouble falling/staying asleep or sleeping too much (8.9%), feeling tired or having little energy (8.8%), feeling nervous (8.2%), trouble relaxing (6.4%), excessive worrying (5.7%) or constant worrying (4.6%).

#### Lifestyle impacts

Since March, the alcohol consumption increased for 1499 (20.8%) of the participants. Among the 544 current smokers (6.9% of the participants), 175 (32.6%) increased their consumption, 60 (11.1%) decreased their consumption whereas 302 (56.3%) did not change. Among the 605 individuals who used cannabis in the past 12 months (7.6% of the participants), 84 (14.2%) increased their consumption, 125 (21.0%) decreased their consumption whereas 385 (64.8%) did not change. The physical activity decreased for 1,757 (22.2%) of the participants. Sleep duration and quality did not change for 7,497 (94.8%) and 7,433 (94.0%), respectively. The food intake increased for 131 (1.7%), while the food quality decreased for 95 (1.2%) of the participants.

Because of the pandemic, 1,355 of the participants (18.6%) looked for support from family, 937 (13.4%) from general media (TV, internet, social media), 890 (12.6%) from the government, 870 (12.4%) from friends, 706 (10.3%) from provincial or federal Health authorities, 485 (7.09%) from professional and 278 (4.14%) from colleagues.

#### Sex differences for early impacts of the COVID-19 outbreak

The COVID-19 outbreak had different impact between men and women. The alcohol consumption increased more among women (22.4% versus 18.5%, *p* < 0.001). Physical activity and quality of sleep decreased more for the women (23.4% versus 20.5%, *p* < 0.001; 5.6% versus 3.3%, *p* < 0.001, respectively). The quantity of food intake increased (2.1% versus 1%, *p* < 0.001) more for the women.

Since March, women accessed more mental health services for anxiety (2.6% versus 1.1%, *p* < 0.001), stress (2% versus 0.7%, *p* < 0.001) and depression (1.6% versus 0.8%, *p* = 0.0033). Nevertheless, the current mental/emotional health was very good or excellent for 74.5% of men but only 65.4% of women (*p* < 0.001). Women have looked more for support from family (22.5% versus 12.9%, *p* < 0.001), general media (16.2% versus 9.6%, *p* < 0.001), friends (15.4% versus 8.1%, *p* < 0.001), provincial/federal Health authorities (11.7% versus 8.3%, *p* < 0.001), professional (8.7% versus 4.9%, *p* < 0.001), colleagues (4.9% versus 3.1%, *p* < 0.001). Looking for support from the government (financial support, financial relief, resources) was not different between women and men.

Over the last 2 weeks, women were more bothered by the following problems, more than half of the days or nearly every day: trouble falling/staying asleep or sleeping too much (10.7% versus 6.2%, *p* < 0.001), feeling tired or having little energy (10.6% versus 6.2%, *p* < 0.001), feeling nervous (10.9% versus 4.4%, *p* < 0.001), trouble relaxing (8.2% versus 3.8%, *p* < 0.001), excessive worrying (7.3% versus 3.2%, *p* < 0.001) or constant worrying (6.1% versus 2.3%, *p* < 0.001).

#### Age differences for early impacts of the COVID-19 outbreak

When looking to the COVID-19 outbreak impact among individuals under and over 65 years, the income substantially decreased for less elderly (7.8% versus 2.6%, *p* < 0.001). The alcohol consumption increased more for the younger (24% versus 15.9%, *p* < 0.001). The quality of sleep and food decreased more for the younger (5.7% versus 3.1%, *p* < 0.001; 1.6% versus 0.6%, *p* < 0.001, respectively). The quantity of intake increased more for the younger (2% versus 1.1%, *p* = 0.0021). The younger accessed more mental health services for anxiety (2.5% versus 1.2%, *p* < 0.001), stress (1.9% versus 0.8%, *p* < 0.001) and depression (1.7% versus 0.6%, *p* < 0.001). Nevertheless, the current mental/emotional health was very good or excellent for 74.3% of elderly but only 65.8% of younger (*p* < 0.001).

Since March, elderly have looked more for support from family (26.2% versus 13.6%, *p* < 0.001), friends (17.4% versus 9.1%, *p* < 0.001), general media (15.8% versus 11.9%, *p* < 0.001), provincial/federal Health authorities (11.6% versus 9.4%, *p* = 0.0037), professional (8.7% versus 6.1%, *p* < 0.001) or community/volunteer organization (4.1% versus 1.8%, *p* < 0.001). Looking for support from colleagues was less frequent among the elderly (2.2% versus 5.3%, *p* < 0.001).

Over the last 2 weeks, the younger were bothered more by the following problems, more than half of the days or nearly every day: trouble falling/staying asleep or sleeping too much (10.4% versus 6.5%, *p* < 0.001), feeling tired or having little energy (10.4% versus 6.3%, *p* < 0.001), feeling nervous (9.8% versus 5.7%, *p* < 0.001), trouble relaxing (7.9% versus 4%, *p* < 0.001), excessive worrying (6.9% versus 3.8%, *p* < 0.001) or constant worrying (5.7% versus 2.8%, *p* < 0.001).

### Association between being tested and demographic, risk exposure, pre-existing medical conditions and COVID-related symptoms

We compared the demographic, exposure and clinical characteristics of the participants who have been tested to those who have not. Participants below the median age were most frequently tested (*p* < 0.001). There was no difference between men and women. Participants living in the Montreal area were more likely to be tested than those living outside of Montreal (9.3% versus 5.2%, *p* < 0.001). Participants living in a house or duplex were less likely to be tested than those living in apartment or condominium (7.3% versus 9.4%, *p* = 0.002). This relationship is still significant when adjusted for living in Montreal or outside. Participants coming back from international travel were most frequently tested (9.8% versus 7.4%, *p* = 0.001). Medical workers and people having contact with a COVID-19 positive individual were most frequently tested (32.0% versus 6.5%, *p* < 0.001 and 42.4% versus 6.1%, *p* < 0.001, respectively).

Participants having at least one pre-existing medical condition (as defined above) were more frequently tested (*p* = 0.002). In particular, individuals having a preexisting condition such as asthma (*p* < 0.001), COPD (*p* < 0.001), chronic bronchitis (*p* < 0.001), nonalcoholic fatty liver disease (NAFLD) (*p* < 0.001) were more frequently tested (Supplementary Table 1).

A multiple logistic regression analyzis showed that place of residence, dweling, risk exposure (medical worker, contact with a COVID-19 positive patient, international travel), having at least one pre-existing condition were independent factors associated with the outcome (Table 1).

**Table 1.** Multivariate analyzis for socio-demographic, medical and exposure factors for being tested

| Variables | OR [CI<sub>95%</sub>] | p value |
| --- | --- | --- |
| Age (years) | 0.99 [0.98-1] | 0.20919 |
| Gender (Ref: female) | 1.12 [0.92-1.36] | 0.25101 |
| Living in Montreal (Ref: other) | 1.79 [1.43-2.25] | <0.0001 |
| Dwelling (Ref: House) | 1.32 [1.07-1.62] | 0.00817 |
| Contact with a COVID-19 case (Ref: No) | 6.57 [4.9-8.76] | <0.0001 |
| Medical worker (Ref: No) | 4.03 [2.99-5.39] | <0.0001 |
| Having at least one medical precondition (Ref: No) | 1.45 [1.19-1.76] | 0.00016 |
| International traveller (Ref: No) | 1.43 [1.15-1.78] | 0.00124 |

All the COVID-related symptoms were more frequent among tested participants (Supplementary Table 1).

Taking into account socio-demographic, medical and exposure factors (place of residence, dweling, medical worker, contact with a COVID-19 positive patient, international travel, pre-existing condition) as confounding factors and COVID-related symptoms as explanatory variables, we performed a GPLTR analyzis for identifying the combinations of symptoms leading to the most homogeneous sub-groups with respect to being tested. The procedure with the BIC criterion led to a final tree having five leaves built with three selected COVID-related symptoms: dyspnea, dry cough and fever (Figure 1A).

**Figure 1.**
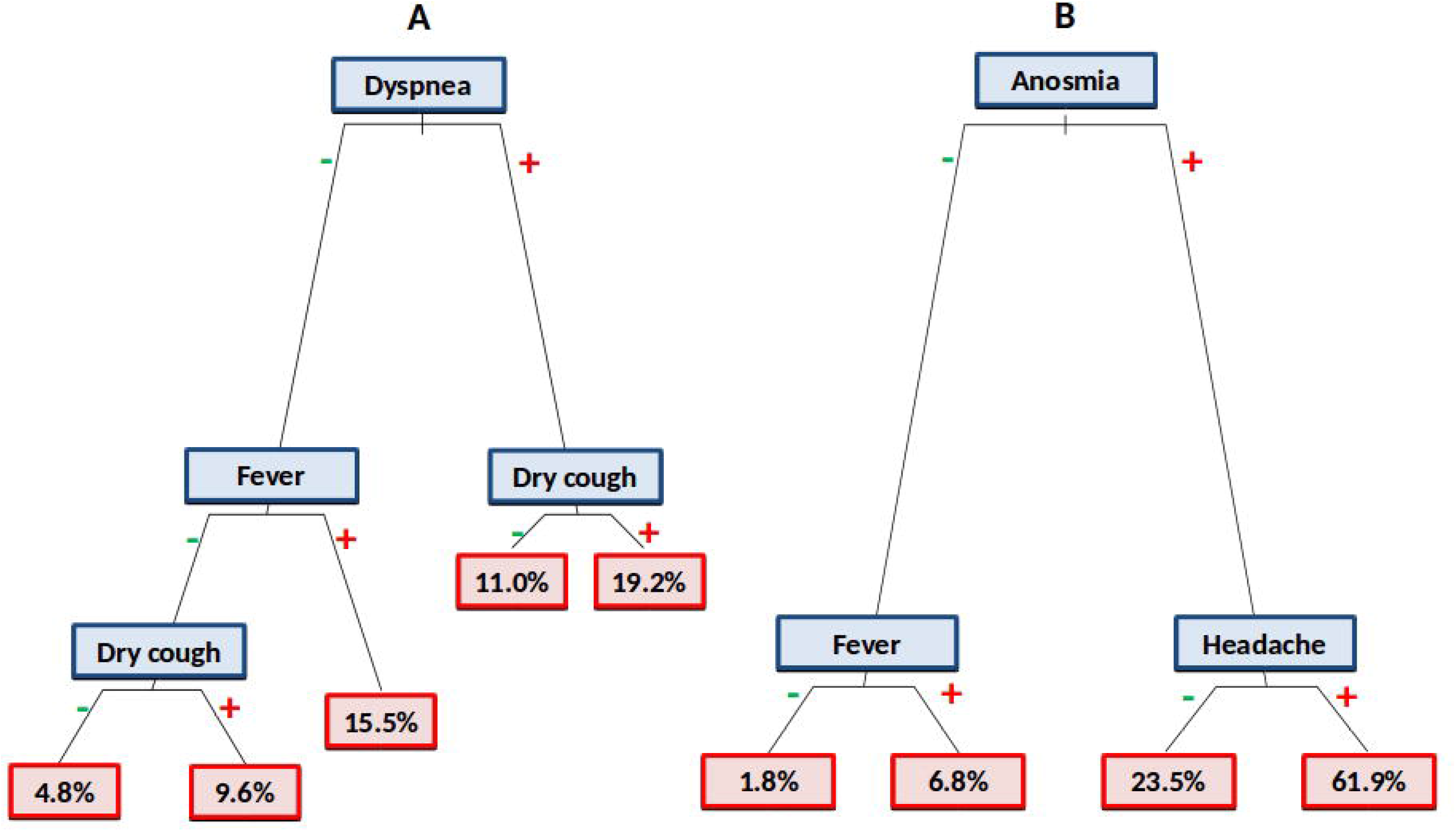
GPLTR model for being tested (A), for being positive when tested (B)

We identified: (i) a leaf with a very low rate for being tested (4.8%, 251/5206) with none of the selected COVID-related symptoms; (ii) a leaf with the highest rate 19.2% (150/780) of tested individuals having both dyspnea and dry cough; (iii) three leaves with at least one of the symptoms with a rate of being tested: dyspnea (11.0%, 55/500), fever (15.5%, 71/459), dry cough (9.6%, 94/978).

Table 2 displays odd-ratio estimates and p-values for the confounding variables and terminal leaves corresponding to the optimal GPLTR tree for being tested.

**Table 2.** GPLTR analyzis for confounding factors and COVID-related symptoms for being tested

| Variables | OR [CI<sub>95%</sub>] | p value |
| --- | --- | --- |
| Living in Montreal (Ref: other) | 1.70 [1.38-2.11] | <0.0001 |
| Dwelling (Ref: House) | 1.36 [1.12-1.65] | 0.001 |
| Contact with a COVID-19 case (Ref: No) | 6.31 [4.71-8.42] | <0.0001 |
| Medical worker (Ref: No) | 4.22 [3.23-5.49] | <0.0001 |
| Having at least one medical precondition (Ref: No) | 1.39 [1.16-1.66] | 0.0003 |
| International traveller (Ref: No) | 1.44 [1.17-1.76] | 0.0004 |
| Wet Cough (Ref: no fever, wet cough, dyspnea) | 1.95 [1.50-2.54] | <0.0001 |
| Fever (Ref: no fever, wet cough, dyspnea) | 3.60 [2.65-4.84] | <0.0001 |
| Dyspnea (Ref: no fever, wet cough, dyspnea) | 2.59 [1.86-3.55] | <0.0001 |
| Dyspnea & Fever (Ref: no fever, wet cough, dyspnea) | 4.49 [3.56-5.66] | <0.0001 |

### Association between being COVID-19 positive and demographic, risk exposure, medical pre-existing conditions and COVID-related symptoms

Participants below the median age were most frequently positive (*p* < 0.001). There were more women among positives (8.2% versus 3.4%, *p* = 0.02) but it was no longer significant after multiple testing adjustment. There was no difference between geographical area of residence or international travelers. Medical workers (*p* < 0.001) and people having contact with a COVID-19 positive individual (*p* < 0.001) were most frequent among positives (Supplementary Table 2). Participants having at least one medical pre-existing condition (as defined above) were less frequently positive (4.3% versus 8.7% *p* = 0.04) but it was no longer significant after multiple testing adjustment.

Other variables such as blood group, treatments, pre-existing conditions, vaccination (Influenza and BCG), smoking, BMI, presence in the household of children or pets were not significantly associated with positive status.

A multiple logistic regression analyzis showed that being a medical worker or having contact with a COVID-19 patient were the only independent factors associated with the outcome (Table 3).

**Table 3.** Multivariate analyzis for socio-demographic and exposure factors for being positive

| Variables | OR [CI<sub>95%</sub>] | p value |
| --- | --- | --- |
| Age (years) | 0.97 [0.91-1.03] | 0.30 |
| Gender (Ref: female) | 0.48 [0.18-1.17] | 0.12 |
| Contact with a COVID-19 case (Ref: No) | 5.29 [2.21-13.37] | <0.0001 |
| Medical worker (Ref: No) | 2.62 [1.08-6.52] | 0.03 |

Patients with loss of taste and smell, fever, dyspnea, headache, pain, fatigue, shivering, loss of appetite, dry cough and nausea symptoms were more frequent among positive individuals. Wet cough and other symptoms were not significantly associated (see Supplementary Table 2).

When estimating the probability of being positive given that the person has experienced a particular symptom (see statistical section), we showed (Figure 2, left panel) that symptoms with the highest probabilities are ageusia, anosmia, loss of appetite, headache, fatigue. In contrast, ear, nose and throat (ENT) symptoms (running nose, sore throat, hoarseness, sinus pain) and wet cough showed the lowest probabilities.

**Figure 2.**
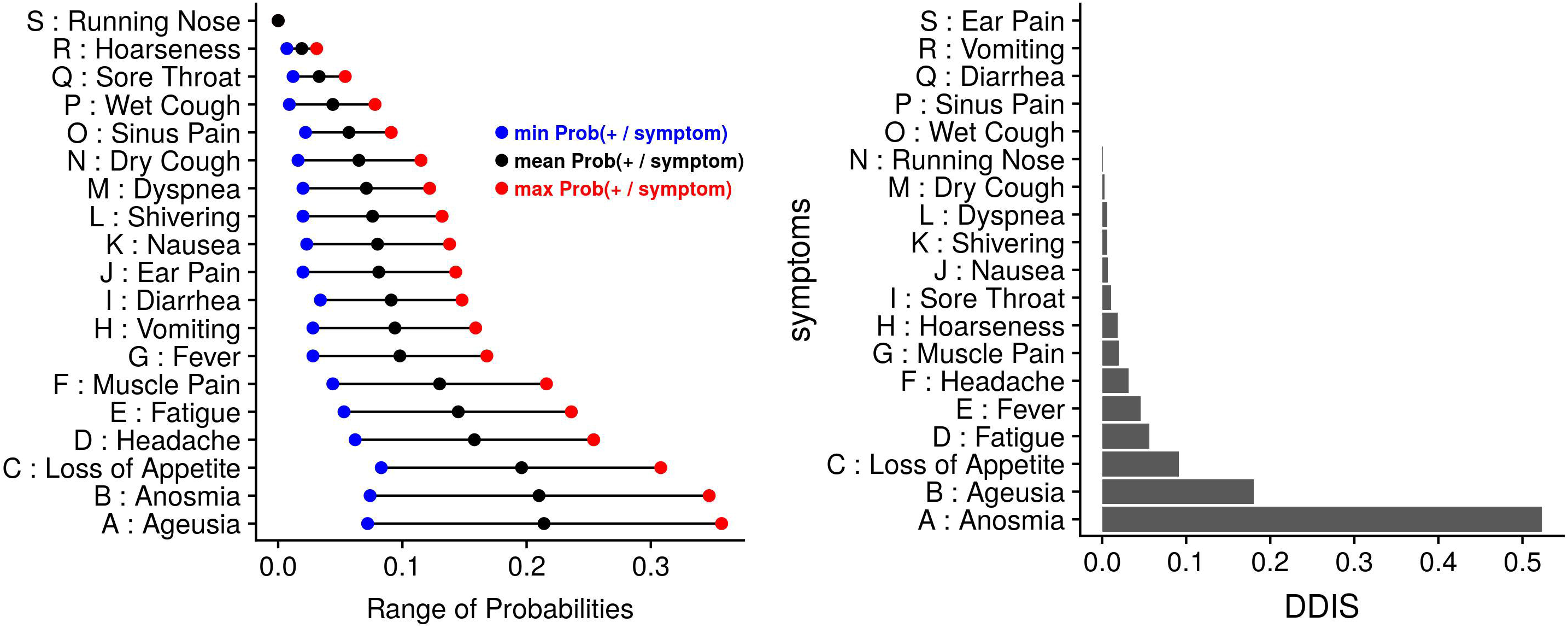
Relationship between symptoms and positivity Left panel: Probability of being positive given that the person has experienced a symptom Right panel: Normalized Depth Deviance Importance Score of the symptoms

Taking into account exposure status (contact with COVID-19 infected patient, medical worker) as confounding factors and COVID-related symptoms as explanatory variables, we performed a GPLTR analyzis for identifying the combinations of symptoms leading to the most homogeneous sub-groups with respect to being positive. Our procedure with the BIC criterion led to a tree with four leaves built with three different COVID-related symptoms selected: anosmia, headache and fever. We identified: (i) a leaf with a very low rate of being positive (1.8%, 8/446) with no anosmia and no fever; (ii) a leaf with a low rate (6.8%, 8/118) of being positive with fever but without anosmia; (iii) a leaf with a moderate rate (23.5%, 12/51) of being positive with anosmia but no headache; (iv) a leaf with a very high positive rate with both anosmia and headache (61.9%, 13/21). Results from the bagging procedure associated with the tree-based model provided a ranking of the variables based on depth deviance importance scores (DDIS). Anosmia, fever and headache selected in the optimal GPLTR tree showed high importance scores (Figure 2, right panel).

Table 4 displays odd-ratio estimates and p-values for the confounding variables and terminal leaves corresponding to the optimal GPLTR tree for positivity.

**Table 4.** GPLTR analyzis for confounding factors and COVID-related symptoms for being positive

| Variables | OR [CI<sub>95%</sub>] | p value |
| --- | --- | --- |
| Contact with a COVID-19 case (Ref: No) | 5.93 [2.42-15.27] | <0.0001 |
| Medical worker (Ref: No) | 4.49 [1.88-11.73] | 0.001 |
| Fever (Ref: no anosmia, fever, headache) | 5.47 [1.90-16.04] | 0.002 |
| Anosmia (Ref: no anosmia, fever, headache) | 26.82 [9.18-85.98] | <0.0001 |
| Anosmia & Headache (Ref: no anosmia, fever, headache) | 161.19 [45.11-660.34] | <0.0001 |

Among the participants tested, the Menni *et al*. predictive model was available for 634 individuals. The model led to an AUC of 83.6% (Supplementary Figure 2). The sensitivity was of 53.7% and the specificity was of 91.4%.

## Discussion

We report the results of an online survey of population-based cohort participants whose main objective was to analyze the characteristics and consequences of the first pandemic wave of COVID-19 spring outbreak in Quebec, Canada. We also report the exposure risk factors and COVID-related symptoms associated with the status of having been tested for SARS-CoV-2 and having been declared positive.

As seen from our analyzis, the demographic characteristics of the respondents are broadly representative of the middle-aged and older population of Quebec. Montrealers, individuals living in an apartment/condominium, having a pre-existing medical condition and having risk exposure (medical worker, contact with a COVID-19 patient, international travel) were more frequently tested. As expected, medical workers and individuals with known or suspected contact with a COVID-positive individual were the most tested. These later findings are in accordance with public health notices giving priority to health care workers and individuals in contact with people tested positive for SARS-Cov-2, whether they had symptoms or not.

Results from the extended tree-based model analyzis, adjusted on exposure factors, show that the combination of dyspnea, dry cough and fever are highly associated with being tested. These results are consistent with the case definition chosen by the Public Health authorities in early spring. The fact that anosmia is not selected reflects its paucity in the general population (4.2% as compared to 11.7% to 25% for the other three symptoms) and that it has been added later in the official list of the main symptoms.

Among the COVID-related symptoms associated with testing, ear, nose and throat (ENT) symptoms (running nose, sore throat, hoarseness, sinus pain) and wet cough were not related to a positive test in univariate analyzis. Results from the extended tree-based model analyzis, adjusted on exposure factors, show that a combination of anosmia, fever and headache are the most discriminant factors for a positive test in our series. Individuals with both anosmia and headache had the highest chance (almost two third) of being positive while those with anosmia alone had almost one-fourth chance of being positive. Individuals without anosmia and fever had less than two percent chance of being positive. These results underline the importance of neurological symptoms such as anosmia and headache, as compared to ENT and gastro-intestinal symptoms. Results obtained from the bagging procedure confirm the importance of the selected symptoms and suggest that the final tree-based model is sufficiently reliable. They also show the importance of fatigue and loss of appetite that are, with anosmia, the main factors of the Menni et al. predictive model. In our series, the operating characteristics of the Menni et al. predictive model are close to those published with a weak sensitivity and a high specificity. However, it is difficult to disentangle the effect of the testing criteria applied in our population from those of the Menni et al. algorithm since the positive cases are only those being tested.

It is worth noting that among the 41 positive individuals, five did not meet the four criteria chosen by the Public Health authorities, two of them reported few symptoms such as fatigue, shivering without fever and/or loss of appetite and three individuals did not experience COVID-related symptoms. Interestingly, all these five patients were tested for having a contact with people being tested positive for SARS-Cov-2. This highlights that a non-negligible fraction of infected people could be asymptomatic or pauci-symptomatic, even in this age group. It underlines the importance of contact tracing as an essential component of the toolbox for containing the COVID-19 outbreak.

Interestingly, we observe some discrepancies between being tested and being positive among the studied factors. Some of them are linked to both outcomes such as being a medical worker, having a contact with a COVID-19 patient and fever. Anosmia increases the discriminative power for being positive as compared to being tested, reflecting its value for positivity. In contrast, dyspnea, that was the main factor for being tested, has a lower discriminative power for positivity.

Some factors associated with testing were not related to a positive test such as international travel, pre-existing conditions and ENT symptoms. While the first COVID-19 cases were linked to international travelers, the public health measure (e.g. quarantine, border closure) mitigated this risk. The lack of relevance for the ENT symptoms should lead to withdraw these symptoms from the list of COVID-related symptoms. For the pre-existing conditions, this discrepancy reflects the testing policies that have focused on group of patients that might be of high risk for severe disease if exposed to the virus. It is worth to note that this over-representation of people having a pre-existing condition led to an inverse relationship with a positive test that is however no longer significant after multiple testing adjustment. Such spurious association could be related to an ascertainment bias caused by testing primarily individuals reporting specific conditions and thought to be positive.

As reported by the participants, the economic consequences in this early stage of the pandemic seem still moderate but is more important for younger participants. However, the real economic impact of the pandemic may appear in the coming months. The alcohol and food consumption slightly increased during the lockdown, especially among women and younger participants. The current emotional health is considered good or excellent for the majority of the participants, but lower for women. As feared, one third of the participants have experienced a decreased access to health services with surgery or medical procedures canceled or deferred for more than ten percent of them. However, it is interesting to note that virtual consultations were widely used.

One of the main strengths of this study is its embedding within the CARTaGENE cohort. It provides a rich body of information collected before the pandemic that is representative of the middle-aged and older population, which seems to be the most commonly affected group. Thus, it offers a unique opportunity to appreciate the impact of COVID-19 infection and public health measures in the Quebec population. In particular, we highlight the selection process for testing at the population level. We show that the most exposed people (medical worker and people having a contact with a COVID-19 positive patient) were more widely tested than the rest of the population. In this later group, only the individuals having all the COVID-related symptoms criteria had a high probability of being tested. This emphasizes the need of increasing the accessibility of testing for the general population who meet the testing criteria through easy to access point-of-care or drive-thru testing centers.

There are however some limitations to this study. Firstly, we experienced a low response rate that might be explained by the online survey in a population that was previously used to paper surveys and by the short time allowed to respond. Nevertheless, our series is broadly representative of the entire cohort. Secondly, it relies on self-reported data, which could be subjected to biases in respondents’ recall and to potential effect from mainstream media coverage. Thirdly, when analyzing the factors related to a positive test, there is an ascertainment bias caused by testing primarily individuals reporting specific exposures or symptoms. This may blur some relationship between factors and a positive outcome. This highlights the interest of seroepidemiological studies at the population level. Fourthly, our population is limited to middle and older adults and has less than one percent of individuals living in senior’s house that were severely affected by the COVID-19 outbreak in the province.

Finally, this study is a first step toward a follow-up study intended to understand the course of the pandemic and its consequences in the population. It will also allow us to compare the public health policies between provinces and countries. Moreover, it will be enriched with a serological study in order to analyze more thoroughly the non-tested symptomatic and asymptomatic cases.

## Conclusion

To our best knowledge, this is one of the first report about the consequences of COVID-19 outbreak at the population level. We have shown some discrepancies between the symptoms and risk factors associated with being tested and being positive. In particular, the anosmia is a major discriminant symptom for positivity whereas ENT symptoms should be withdrawn from the list of COVID-related symptoms. It also emphasizes the need of increasing the accessibility of testing for the general population.

## Data Availability

The data that support the findings of this study are available from CARTaGENE but restrictions apply to the availability of these data. Data are available directly from CARTaGENE (http://cartagene.qc.ca; +1 514-345-2156).

## Author’s contributions

RJ: conceptualization, data curation, formal analyzis, investigation, methodology, writing - original draft. NN: conceptualization, resources, writing - original draf. SCB: writing - original draft. CL: resources. TM: data curation, software. YP: data curation, software. PB: conceptualization, formal analyzis, methodology, project administration, supervision, validation, writing - original draft.

## Consent for publication

Not applicable

## Ethics approval and consent to participate

This project has been approved by the Research Ethics Board of the CHU Sainte-justine on 28th May 2020 under the reference MP-21-2011-345, 3297. All participants have given their consent.

## Acknowledgements

We would like to thank all the CARTaGENE participants for their generous investments in health research. We would also like to thank the RAMQ and the Commission d’acces a l’information (CAI) for their support in obtaining the data.

## Availability of data and materials

Tables

**Additional Files**

Additional file 1 — supplementary.odt

Tables S1 and S2.

Additional file 2 — Fig_S1.tif

Figure S1: Number of tested participants and cumulative number of tested participants over time.

